# Background prostate tissue is quantitatively abnormal on MRI in patients with clinically significant prostate cancer

**DOI:** 10.1101/2022.10.12.22280855

**Authors:** Christopher C Conlin, Roshan Karunamuni, Troy S Hussain, Allison Y Zhong, Karoline Kallis, Deondre D Do, Asona J Lui, Garnier Mani, Courtney Ollison, Mariluz Rojo Domingo, Ahmed Shabaik, Christopher J Kane, Aditya Bagrodia, Rana R McKay, Joshua M Kuperman, Rebecca Rakow-Penner, Michael E Hahn, Anders M Dale, Tyler M Seibert

## Abstract

**Background:** *T*_*2*_-weighted MRI is standard for detecting clinically significant prostate cancer (csPCa) by identifying visible lesions that stand out from the background prostate.

**Purpose:** To determine whether patients with csPCa have abnormal *T*_*2*_-weighted signal in non-lesion, background prostate tissue (BP).

**Methods:** This retrospective study included two patient cohorts who underwent 3T MRI examination for suspected csPCa. Median (urine-normalized) *T*_*2*_-weighted signal was computed for BP and compared between patients with and without csPCa. csPCa discrimination performance of *T*_*2*_-weighted BP signal was evaluated using area under receiver operating characteristic curves (AUC). *T*_*2*_ and *S*_*0*_ (a proxy for proton density) were computed and compared between patients with and without csPCa. *T*_*2*_ was also recomputed using larger buffers around csPCa lesions. csPCa discrimination performance was compared between two predictors: Restriction Spectrum Imaging (RSI) *C*_*1*_ and RSI *C*_*1*_ normalized by global prostate median *T*_*2*_-weighted signal.

**Results:** Cohort 1: 46 patients (age: 64±10 years). Cohort 2: 151 patients (65±8 years). Urine-normalized *T*_*2*_-weighted signal was systematically lower in BP of subjects with csPCa (*p*≤0.034) and indicated the presence of cancer (cohort 1: AUC=0.80; cohort 2: AUC=0.68). BP *T*_*2*_ was significantly lower in csPCa patients (*p*≤0.011), while *S*_*0*_ was not (*p*≥0.30). BP *T*_*2*_ measurements were stable to within 5% with buffers from 0 to 30 mm around visible lesions. csPCa discrimination improved with incorporation of BP *T*_*2*_-weighted signal (cohort 1: AUC=0.72 for RSI *C*_*1*_ alone, versus 0.81 with BP *T*_*2*_-weighted signal; cohort 2: AUC=0.63 versus 0.76).

**Conclusion:** Lower *T*_*2*_-weighted signal in BP suggests the presence of csPCa.

## Introduction

While many studies have established the value of *T*_*2*_ for detection of clinically significant prostate cancer (csPCa) (1–6), these have focused on characterizing the *T*_*2*_ within radiographically visible lesions. Current practice, per standardized reporting guidelines (PI-RADS), is to compare local *T*_*2*_-weighted signal to the background signal within the prostate to identify hypointense lesions that may represent csPCa. However, there are several common processes (MRI-invisible cancer, pre-cancerous lesions, inflammation, some components of benign prostatic hyperplasia, etc.) that can lead to darker *T*_*2*_-weighted signal throughout the gland (7–9), thus potentially undermining the contrast of csPCa compared to the background. The present study seeks to determine whether patients with csPCa have abnormal *T*_*2*_-weighted signal in prostate tissue *outside* of the index lesions identified on MRI—i.e., in the background prostate (BP).

It has long been known that csPCa arises within a field effect of histopathological abnormalities (10,11). More recently, genomic studies have demonstrated that even the benign BP in patients with csPCa has a wide range of genetic alterations (12). It is unclear to what extent BP is changed because of germline features, somatic mutations, inflammatory changes, and/or reactive processes secondary to the presence of csPCa in the gland. Identifying patients with abnormal MRI features outside visible lesions might provide insight into underlying biology. BP abnormalities could also complement lesion-level features in estimating the probability of a patient having csPCa and therefore potentially useful in deciding which patients need to undergo prostate biopsy.

Here, we test two independent patient cohorts for a systematic decrease in the *T*_*2*_-weighted signal of BP in patients with csPCa. We hypothesize that *T*_*2*_-weighted signal from BP can indicate the presence of csPCa. We then examine whether *T*_*2*_ or proton density effects are driving the observed reduction in *T*_*2*_-weighted signal, and whether these effects stem from normal age-related changes to the prostate or simply from cancer adjacent to radiological lesions (8). Finally, we test whether *T*_*2*_-weighted signal characteristics of BP provide information complementary to focal restricted diffusion-weighted MRI (DWI) to improve patient-level detection of csPCa.

## Materials and Methods

### Study population

This retrospective study was approved by the local institutional review board (IRB). A waiver of consent was obtained from the IRB to access patient MRI data and other clinical records. Two cohorts of patients with suspected csPCa were included in this study, independent in time and in MRI acquisition protocol. The first cohort was comprised of 81 consecutive men who underwent MRI examination on a single scanner between August and December of 2016. The second cohort included consecutive 440 men who underwent MRI examination on the same scanner between January of 2017 and February of 2020, after implementation of a change in acquisition protocol. Patients were included in analyses if they had no prior treatment for prostate cancer; in the larger cohort 2, patients were additionally excluded if they did not have a biopsy within 6 months of the MRI. Both cohorts have been studied previously for characterization of signal in lesions (13–15). BP signal has not been reported previously.

### MRI acquisition

All MR imaging was performed on 3T clinical scanners (Discovery MR750; GE Healthcare, Waukesha, WI), using a 32-channel phased-array body coil surrounding the pelvis. Acquisition details are summarized in Table 1. For each patient of cohort 1, two axial DWI volumes were separately acquired using different echo times (TEs) but with other parameters held constant. For cohort 2, a single axial DWI volume was acquired for each patient. For anatomical reference, a higher resolution *T*_*2*_-weighted volume was acquired for all patients, with scan coverage identical to that of the multi-shell DWI volume.

**Table 1:** MRI acquisition details for the two patient cohorts included in this study. DWI volumes were acquired using a diffusion-weighted spin-echo pulse sequence (default tensor) with an echo-planar imaging (EPI) readout. *T*_*2*_-weighted (T2W) volumes were acquired using a fast spin-echo (FSE) pulse sequence. In-phase and out-of-phase *T*_*1*_-weighted images were acquired using “Liver Acquisition with Volume Acquisition (LAVA)-Flex,” a dual-echo, fast spoiled gradient-echo sequence. Water-only and fat-only images were generated automatically from the in-phase/out-of-phase images by scanner software using the 2-point Dixon method. Dynamic contrast-enhanced (DCE) MRI was performed using a time-resolved imaging of contrast kinetics (TRICKS) protocol with a temporal resolution of one frame every 7 seconds. Baseline-subtracted volumes were automatically generated from TRICKS data by the scanner software.

|  |  | FOV<br>(mm) | Matrix<br>[resampled dimensions] | Slices | Slice<br>thickness<br>(mm) | TR<br>(ms) | TE (ms) | b-values (s/mm<sup>2</sup>)<br>[number of samples] |
| --- | --- | --- | --- | --- | --- | --- | --- | --- |
| Cohort 1 | DWI 1 | 220×220 | 96×96 [128×128] | 34 | 3 | 5000 | 80 | 0 [7*], 200 [6], 1000 [6],<br>2000 [6], 3000 [6] |
|  | DWI 2 | 220×220 | 96×96 [128×128] | 34 | 3 | 5000 | 100 | 0 [7*], 200 [6], 1000 [6],<br>2000 [6], 3000 [6] |
|  | T2W | 220×220 | 320×320 [512×512] | 34 | 3 | 6225 | 100 | N/A |
|  | T1<br>LAVA-Flex | 320×256 | 320×224 [512×512] | 140 | 4 | 4 | 2 (in-phase)<br>1 (out-of-phase) | N/A |
|  | DCE | 220×220 | 256×168 [512×512] | 32 | 3 | 4 | 2 | N/A |
| Cohort 2 | DWI | 240×120 | 96×48 [128×64] | 16 | 6 | 4500 | 68 | 0 [2*], 500 [6], 1000 [6],<br>2000 [12] |
|  | T2W | 240×240 | 320×320 [512×512] | 32 | 3 | 6080 | 102 | N/A |
|  | T1<br>LAVA-Flex | 340×272 | 320×224 [512×512] | 140 | 4 | 4 | 2 (in-phase)<br>1 (out-of-phase) | N/A |
|  | DCE | 240×240 | 256×168 [512×512] | 64 | 3 | 4 | 2 | N/A |

\*An extra $b=0$ s/mm<sup>2</sup> volume was acquired with reverse phase encoding to enable correction of $B_0$ -inhomogeneity distortions

### MRI data post-processing

Post-processing and analysis of all MRI data was performed using MATLAB (The MathWorks, Inc; Natick, MA). Diffusion data were first corrected for distortions due to *B*_*0*_ inhomogeneity, gradient nonlinearity, and eddy currents (16–18). The multiple DWI samples acquired at each *b*-value were averaged together. To correct for patient motion between the two separately-acquired DWI volumes of cohort 1, multiscale image registration by intensity correlation was applied (19). To account for arbitrary signal-intensity scaling between acquisitions, all DWI volumes were normalized by the median signal intensity of urine in the bladder at *b*=0 s/mm^2^ (20).

Quantitative *T*_*2*_ mapping was performed for all patients of cohort 1 by fitting the signal values from the two *b*=0 s/mm^2^ volumes acquired at different TEs with the *T*_*2*_-weighted signal decay formula: 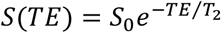, where *S*(*TE*) is the signal measured at a particular TE, *S*_0_ is the signal potential at magnetic equilibrium (which is proportional to proton density (21)), and *T*_*2*_ is the transverse relaxation time. Voxel-wise estimates of both *T*_*2*_ and *S*_*0*_ were recorded for each patient.

For all patients, regions of interest (ROIs) were defined for the whole prostate (WP), peripheral zone (PZ), and transition zone (TZ). Anatomic segmentation/contouring was performed by a radiation oncologist (3 years of experience) and two board-certified radiologists (3 and 6 years of experience, respectively) using MIM software (MIM Software, Inc; Cleveland, OH). Lesions were identified by board-certified radiologists, per clinical routine, using PI-RADS criteria. Lesion ROIs were defined for this study by consensus by two board-certified radiologists using multiparametric MRI (mpMRI) and PI-RADS v2.1. The presence or absence of csPCa (grade group ≥2) was per clinical diagnostic routine using biopsy or prostatectomy specimens.

### Examination of *T*_*2*_-weighted signal in BP

The median signal value on the urine-normalized *b*=0 s/mm^2^ (i.e., *T*_*2*_-weighted) volumes was computed for all ROIs (WP, PZ, and TZ) in BP. For patients without identifiable lesions (whether or not cancer was found on systematic biopsy), this amounted to calculating the median signal within the whole ROI. For patients with visible lesions, the lesion ROI was excluded from the calculation, along with an added 5-mm margin around the lesion ROI to account for the possibility of cancer outside the defined lesion ROI. Formally, the median was computed from all voxels in the set *A* ∩ *B*^*C*^, where *A* is the set of voxels in the gland ROI (WP, PZ, or TZ), *B* is the set of voxels within the lesion ROI dilated by 5 mm, and *B*^*C*^ denotes the complement of *B*. Median BP signal values were compared between patients with csPCa and those without csPCa using two-sample *t*-tests to assess statistical significance (α=0.05). Median signal values were also analogously compared between benign and cancerous lesions.

To examine the diagnostic utility of *T*_*2*_-weighted signal in BP, receiver operating characteristic (ROC) curves were generated at the patient level, using median BP signal values as predictor variables to determine the presence or absence of csPCa on clinical biopsy (generally 12 systematic cores, plus targeted cores for lesions identified on MRI). ROC curves were generated for the median BP signal of each zonal ROI (WP, PZ, and TZ), as well as for the median signal of the whole gland with lesions included. The area under each ROC curve (AUC) was computed to evaluate csPCa discrimination performance.

### Potential causes of BP signal differences between patients with and without csPCa

*T*_*2*_-weighted signal intensity is governed by hardware factors (e.g., receiver gain) and two tissue-specific parameters: proton density (directly related to the signal potential at magnetic equilibrium, *S*_*0*_) and transverse relaxation time (*T*_*2*_) (21). Imaging hardware was consistent for all patients in this study, so any observed differences in *T*_*2*_-weighted BP signal between subjects was assumed to arise from differences in either the proton density or *T*_*2*_ of the BP.

Median *T*_*2*_ (in milliseconds) was computed from the voxel-wise *T*_*2*_ maps of cohort 1 within all BP ROIs to quantify the impact of *T*_*2*_ on BP signal. Similarly, median *S*_*0*_ was computed from the voxel-wise *S*_*0*_ maps of cohort 1 within all BP ROIs to quantify the impact of proton density on BP signal. *T*_*2*_ and *S*_*0*_ measurements from BP were compared between patients with and without csPCa, using two-sample *t*-tests (α=0.05). *T*_*2*_ and *S*_*0*_ values were similarly compared between benign and cancerous lesions.

To evaluate the extent to which age-related effects might be driving any observed differences in BP characteristics between patients with and without csPCa, the Pearson correlation was computed between patient age and any BP parameters (median *T*_*2*_, *S*_*0*_) that were significantly different between patients with and without csPCa.

To ensure that any differences in the BP signal of patients with csPCa were not simply the result of adjacent tumor tissue that was missed during lesion contouring and erroneously included as part of the background prostate, median *T*_*2*_ in the BP was computed multiple times with increasingly large buffers of excluded voxels around the csPCa lesion ROI. Specifically, the median *T*_*2*_ was computed from all voxels in the set *A* ∩ (*B* + *m*)^*C*^, where *A* is the set of voxels in the gland ROI (WP, PZ, or TZ), *B* is the radiographically defined lesion ROI, and *m* is the buffer of excluded voxels around *B*. The width of the buffer *m* around *B* was increased from 0 to 30 mm in increments of one voxel-width, approximately 2.5 mm. For comparison, median *T*_*2*_ was also computed for the whole gland with lesions included.

### Comparison of *T*_*2*_-weighted signal and diffusion in the prostate

To examine if the *T*_*2*_-weighted signal characteristics of the prostate provide additional diagnostic information beyond that provided by patient-level DWI alone, *T*_*2*_-weighted signal in the whole prostate was compared against a previously validated biomarker of prostate cancer based on focal diffusion restriction: Restriction Spectrum Imaging (RSI) *C*_*1*_ (RSI *C*_*1*_). RSI *C*_*1*_ signal was computed from the DWI data of each patient by fitting with a previously described 4-compartment RSI model (13,14). Prior studies have employed the maximum *C*_*1*_ value within the prostate as a diffusion-based indicator of prostate cancer (14,15), so the maximum *C*_*1*_ value within the prostate was recorded for each patient in the present study. The Pearson correlation was then computed between the median *T*_*2*_-weighted signal and maximum *C*_*1*_ value in the prostate. Finally, two ROC curves were generated for each patient cohort, one using only the maximum RSI *C*_*1*_ as the predictor variable, and one using the maximum RSI *C*_*1*_ normalized by the median *T*_*2*_-weighted signal, to indicate the presence or absence of csPCa (grade group ≥2) on clinical biopsy (generally 12 systematic cores, plus targeted cores for lesions identified on MRI).

### Data Availability

The data generated in this study are available upon request from the corresponding author.

## Results

### Study population

A flowchart illustrating patient selection for this study is shown in Figure 1. From cohort 1, 46 patients (age: 64±10 years; PSA: 10.8±17.2 ng/mL) were included. From cohort 2, 151 patients were included (age: 65±8 years; PSA: 11.8±13.9 ng/mL). Radiologic and pathologic characteristics are summarized in Table 2 for both patient cohorts.

**Figure 1:**
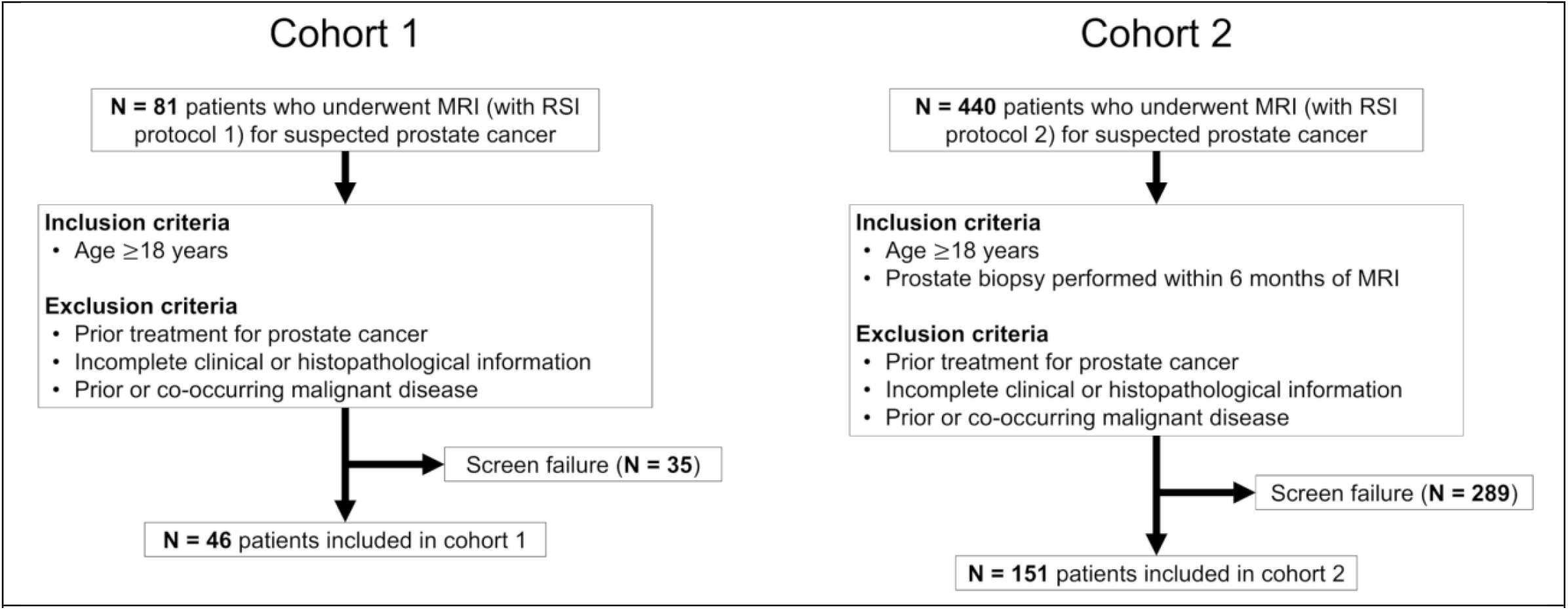
Flowchart illustrating patient selection for both cohorts considered for this study.

**Table 2:** Summary of radiologic and pathologic characteristics of the two patient cohorts included in this study.

|  |  | **Cohort 1** | **Cohort 2** |
| --- | --- | --- | --- |
| **Previous biopsy** | Biopsy naïve | 38 | 103 |
|  | Past biopsy | 8 | 48 |
| **Available pathology** | Systematic biopsy only | 28 | 7 |
|  | Targeted biopsy only | 0 | 17 |
|  | Systematic and targeted biopsy | 15 | 85 |
|  | No biopsy* | 3 | 0 |
|  | Prostatectomy | 12 | 42 |
| **PI-RADS score<sup>†</sup>** | 1 | 10 | 0 |
|  | 2 | 2 | 5 |
|  | 3 | 10 | 27 |
|  | 4 | 10 | 55 |
|  | 5 | 14 | 64 |
| **Gleason Grade Group** | None | 23 | 29 |
|  | 1 | 3 | 39 |
|  | 2 | 8 | 35 |
|  | 3 | 7 | 20 |
|  | 4 | 1 | 16 |
|  | 5 | 4 | 12 |

\*In the smaller Cohort 1, three patients did not have biopsy but had normal MRI and low clinical suspicion of csPCa.
<sup>†</sup>PI-RADS v2 was used for cohort 1, PI-RADS v2.1 for cohort 2

### *T*_*2*_-weighted signal in BP

Figure 2 summarizes the *T*_*2*_-weighted signal characteristics of prostate tissue from both cohorts. Median urine-normalized *T*_*2*_-weighted signal was systematically lower in subjects with csPCa compared to those without, even in BP. In cohort 1, the observed decrease in median *T*_*2*_-weighted signal was statistically significant for each of the examined anatomical zones, with *p*=2.5e-4 for WP, *p*=6.8e-4 for PZ, and *p*=1.3e-4 for TZ. Median *T*_*2*_-weighted signal within csPCa lesions was also significantly lower (*p*=0.003) compared to benign lesions. In cohort 2, the decrease in median *T*_*2*_-weighted signal was significant for all zones (*p*=8.0e-5 for WP, *p*=7.6e-4 for PZ, and *p*=2.0e-4 for TZ), as well as for csPCa lesions compared to benign or clinically insignificant (grade group 1) prostate cancer lesions (*p*=0.034).

**Figure 2:**
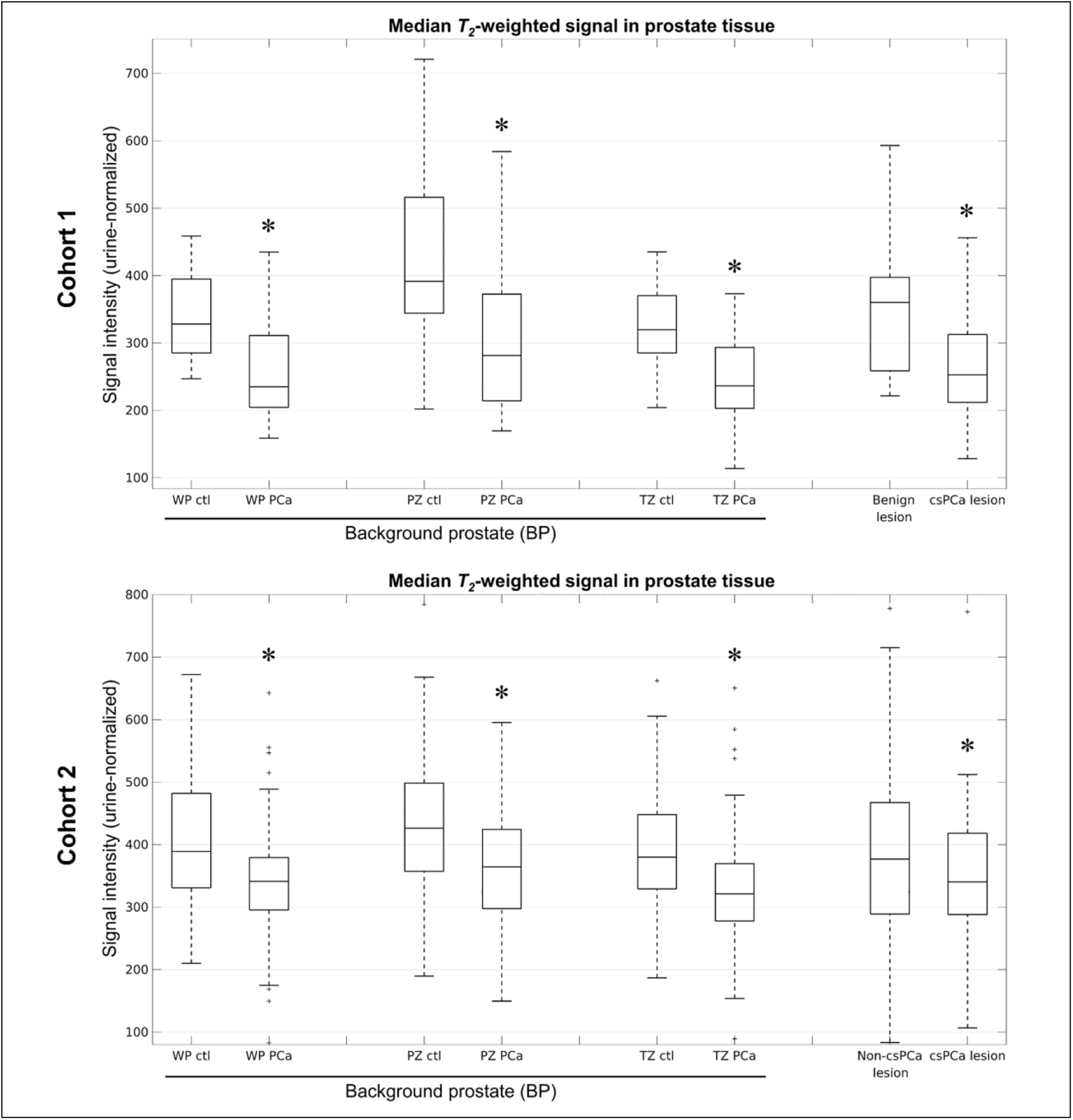
*T*_*2*_-weighted signal in prostate tissue of subjects with and without clinically significant prostate cancer (csPCa). Median (urine-normalized) *T*_*2*_-weighted signal was systematically lower in subjects with csPCa, even in background prostate (BP). Asterisks indicate a significant (*p*<0.05) decrease in the median *T*_*2*_-weighted signal of patients with csPCa compared to those without. WP: whole-prostate, PZ: peripheral zone, TZ: transition zone, ctl: control subject without csPCa, PCa: subject with csPCa. In cohort 2, the “Non-csPCa lesion” group is comprised of subjects with either a benign or clinically insignificant (grade group 1) prostate cancer lesion.

The csPCa discrimination performance of median *T*_*2*_-weighted BP signal is quantified by the ROC curves in Figure 3. In cohort 1, csPCa discrimination performance was similar for BP measurements within the WP and TZ, each having an AUC value of 0.80. The AUC value for BP PZ was slightly lower: 0.77. AUC values for cohort 2 were generally lower than for cohort 1, but still indicated good classification performance of BP signal: 0.68 for WP, 0.66 for PZ, and 0.69 for TZ. In both cohorts, WP classification performance of BP only was nearly identical to the performance with lesions included. The AUCs for BP WP and WP (including lesions) were both 0.80 in cohort 1 and 0.68 in cohort 2.

**Figure 3:**
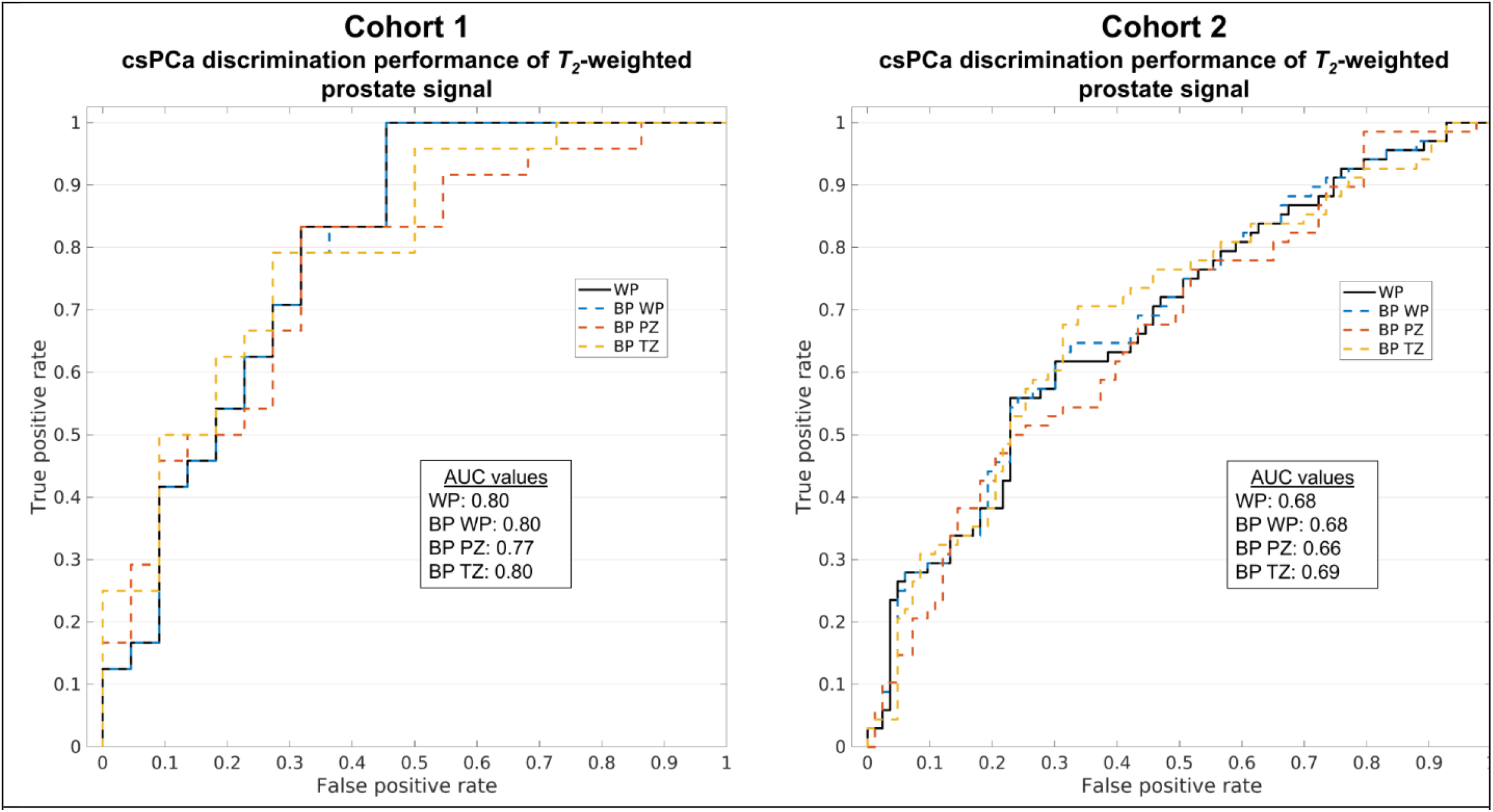
Receiver operating characteristic (ROC) curves illustrating the clinically significant prostate cancer (csPCa) discrimination performance of median (urine-normalized) *T*_*2*_-weighted signal in background prostate (BP). The ROC curve for the whole prostate, including lesions, is shown for comparison (WP). AUC: area under the ROC curve, WP: whole-prostate, PZ: peripheral zone, TZ: transition zone.

### Potential causes of BP signal differences between patients with and without csPCa

Figure 4 summarizes the *T*_*2*_ time and *S*_*0*_ characteristics of prostate tissue from cohort 1 (the only cohort for which multi-TE acquisitions were available to examine these parameters). Median *T*_*2*_ was significantly lower in BP of patients with csPCa compared to those without, with *p*=0.002 for WP, *p*=2.7e-4 for PZ, and *p*=0.011 for TZ. Median *T*_*2*_ was also significantly lower in csPCa lesions compared to benign lesions (*p*=0.009). Median *S*_*0*_, conversely, was not significantly different in patients with csPCa compared to those without, either in BP (*p*=0.30 for WP, *p*=0.39 for PZ, *p*=0.33 for TZ) or lesions (*p*=0.30).

**Figure 4:**
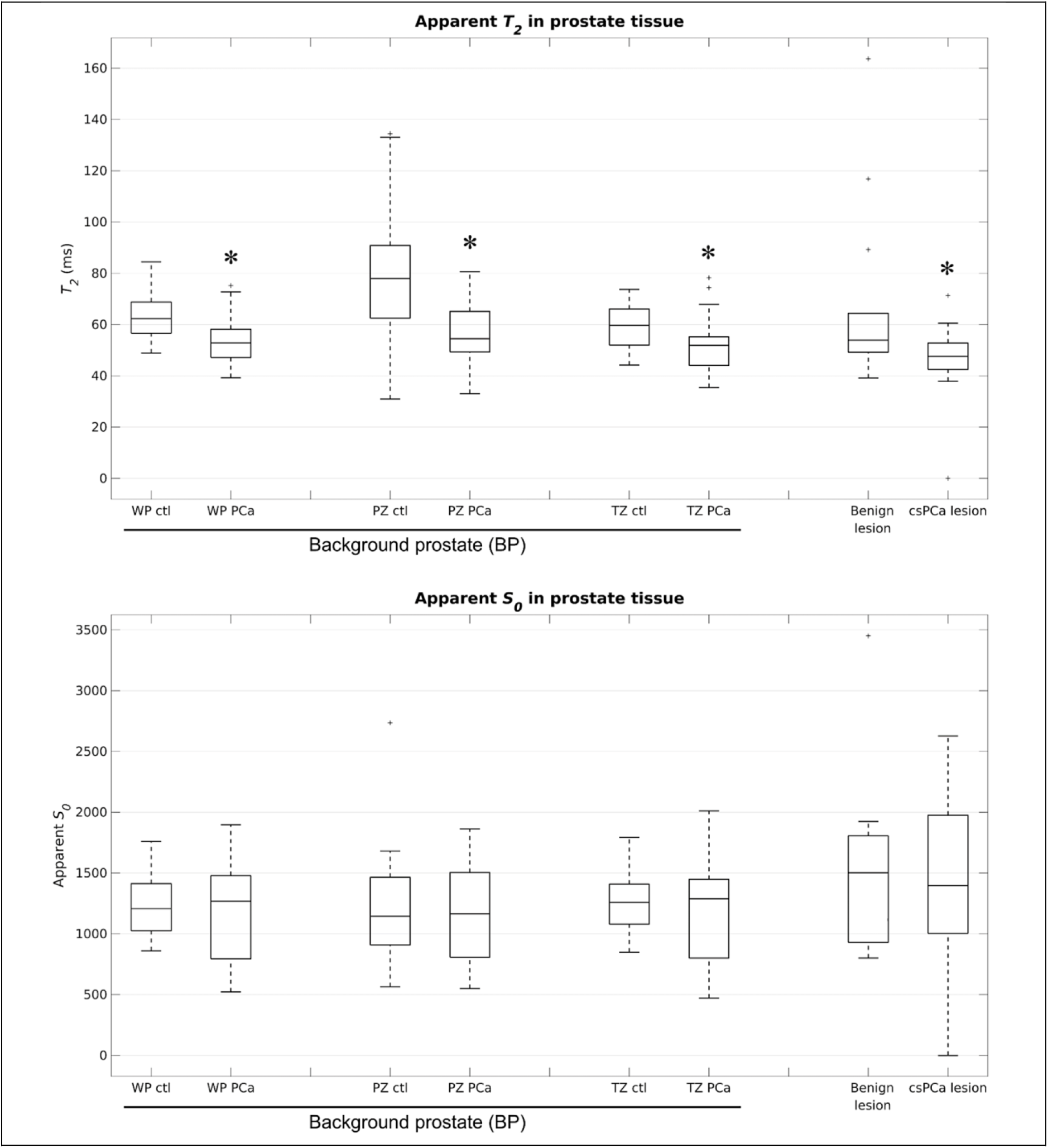
*T*_*2*_ time and *S*_*0*_ in prostate tissue of subjects from cohort 1 with and without clinically significant prostate cancer (csPCa). Asterisks indicate a significant (*p*<0.05) decrease in the measured value from patients with csPCa compared to those without.

The *T*_*2*_ of BP was not significantly correlated with patient age (Supplemental Figure 1), whether measured in the WP (Pearson *r*=-0.02, *p*=0.91), PZ (Pearson *r*=-0.15, *p*=0.32), or TZ (Pearson *r*=0.16, *p*=0.30).

BP *T*_*2*_ measurements from patients with csPCa were relatively insensitive to changes in the size of the margin around the csPCa lesion ROI that was excluded from the BP *T*_*2*_ calculation (Figure 5). BP *T*_*2*_ values were stable within 5% for margin values 0 to 30 mm. BP *T*_*2*_ measurements changed by a maximum of 4.8% in the WP, 4.8% in the PZ, and -4.1% in the TZ. With lesions included in the measurement, WP and TZ *T*_*2*_ values were 0.7% lower than the values measured from BP only. PZ *T*_*2*_ was 4.1% lower with lesions included.

**Figure 5:**
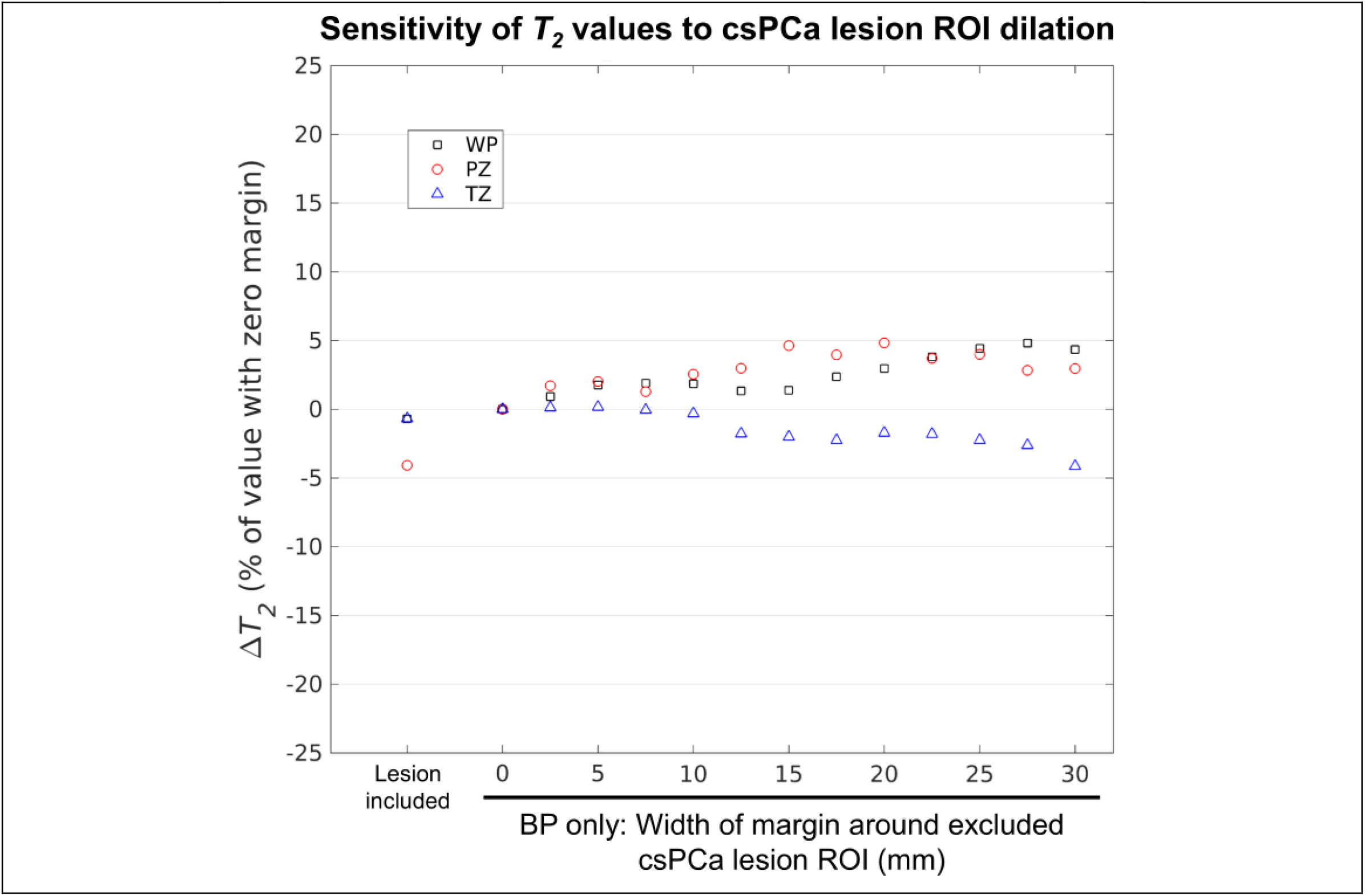
Sensitivity of BP *T*_*2*_ measurements from cohort 1 to the width of the margin around the clinically significant prostate cancer (csPCa) lesion ROI that was excluded from the measurement. For margin widths ranging from 0 to 30 mm, BP *T*_*2*_ measurements were stable to within 5%. BP *T*_*2*_ changed by a maximum of 4.8% in the WP, 4.8% in the PZ, and -4.1% in the TZ. Absolute BP *T*_*2*_ values for a margin width of zero were 52 ms for the WP, 54 ms for the PZ, and 52 ms for the TZ. *T*_*2*_ measurements that included the lesion ROIs are shown for comparison.

### Comparison of *T*_*2*_-weighted signal and RSI *C*_*1*_ in the whole prostate

Supplemental Figure 2 plots whole-prostate measures of maximum RSI *C*_*1*_ (previously shown to be predictive of patient-level csPCa) against median urine-normalized *T*_*2*_-weighted signal. No significant correlation was observed between the two metrics in either cohort (cohort 1: Pearson *r*=-0.01, *p*=0.96; cohort 2: Pearson *r*=0.08, *p*=0.36). Consideration of *T*_*2*_-weighted signal along with RSI *C*_*1*_ yielded improved cancer discrimination performance compared to RSI *C*_*1*_ alone (Figure 6). In cohort 1, the AUC increased from 0.72 for maximum RSI *C*_*1*_ alone to 0.81 for maximum RSI *C*_*1*_ normalized by median *T*_*2*_-weighted signal. In cohort 2, the AUC increased from 0.63 to 0.76.

**Figure 6:**
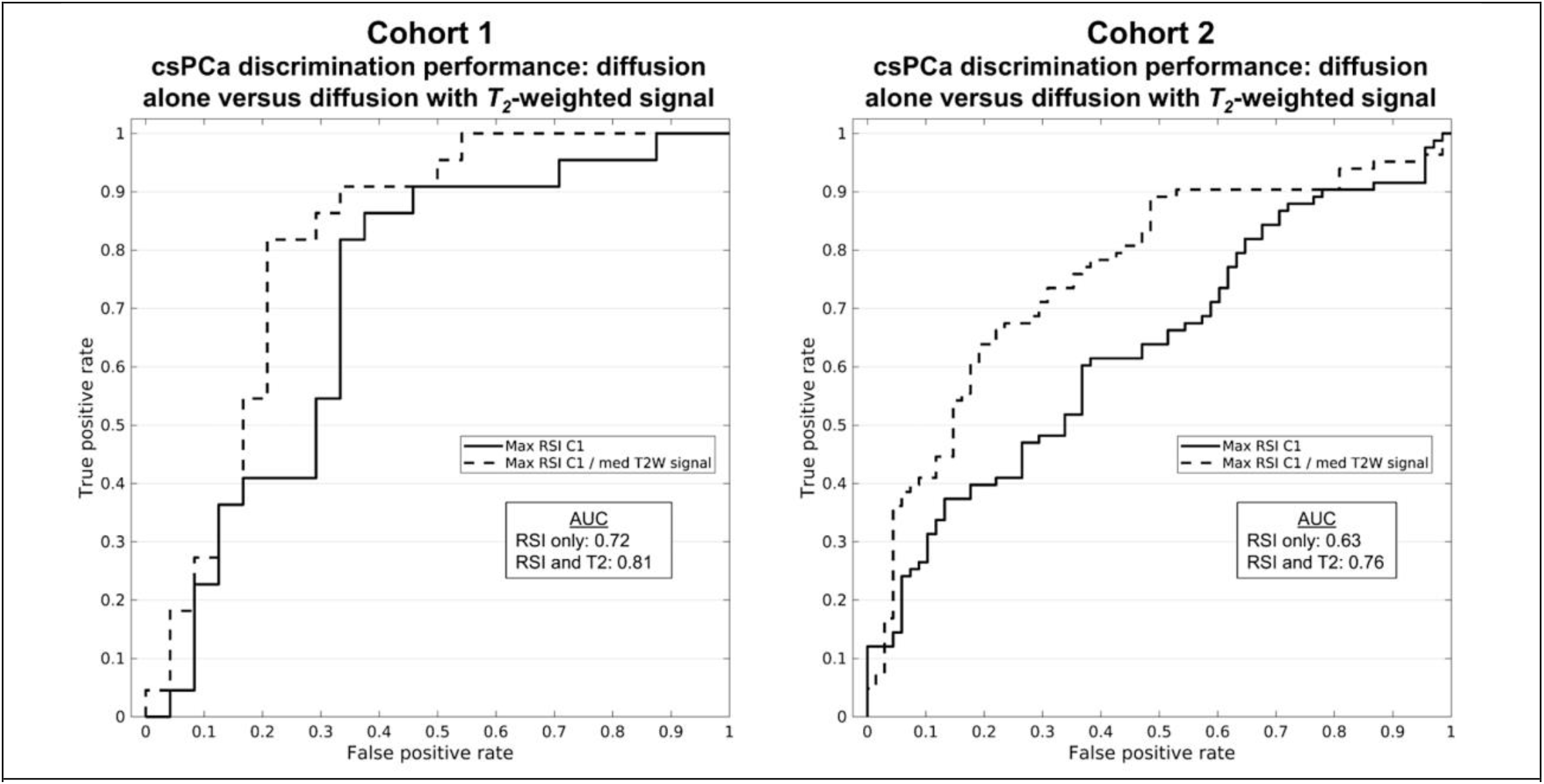
Receiver operating characteristic (ROC) curves illustrating the clinically significant prostate cancer (csPCa) discrimination performance of whole-prostate RSI *C*_*1*_ alone versus whole-prostate RSI *C*_*1*_ normalized by median *T*_*2*_-weighted (T2W) signal. AUC: area under the ROC curve.

## Discussion

To our knowledge, this is the first study to specifically examine *background* prostate tissue for abnormal *T*_*2*_ relaxation associated with the presence of csPCa in the prostate. We found that *T*_*2*_-weighted signal intensity in the BP of patients with csPCa was systematically lower than in patients without csPCa. This systematic decrease in *T*_*2*_-weighted BP signal was observed across two different patient cohorts that were independently acquired using different imaging protocols, suggesting that it is not an artifact of experimental design. Furthermore, *T*_*2*_-weighted signal in the BP meaningfully discriminated patients with or without csPCa, perhaps surprising considering current clinical practice focuses exclusively on suspicious lesions. The csPCa discrimination performance achieved using *T*_*2*_-weighted signal in the BP alone was comparable to the performance reported for methods that examine signal properties within radiographically identified lesions (22–24).

We considered a few explanations for cancer-associated *T*_*2*_-weighted signals observed in BP in both cohorts. First, we noted that *T*_*2*_-weighted signals could be driven by differences in proton density, but multi-TE data from cohort 1 revealed no association of *S*_*0*_ in BP with csPCa. Meanwhile, there was a significant decrease in the apparent *T*_*2*_ time of the BP in patients with csPCa compared to those without. Second, normal age-related changes in prostate *T*_*2*_ have been described (25), but we found no correlation here between BP *T*_*2*_ and patient age. Third, we know that csPCa can be present adjacent to visible lesions (8). Though the median *T*_*2*_ should not be influenced too heavily by inclusion of a relatively small tumor in the BP ROI, we computed the BP *T*_*2*_ value multiple times with increasingly large buffers of excluded voxels around the lesion ROI. If tumor tissue adjacent to the prescribed lesion contour were driving the observed decrease in median BP *T*_*2*_, we would expect that increasing the buffer around the contour would significantly increase the recorded BP *T*_*2*_ values and eliminate the systematic difference between patients with csPCa and those without. However, we found the median BP *T*_*2*_ to be largely insensitive to the lesion buffer size, making lesion-adjacent cancer an unlikely explanation for BP signal differences.

Patients with csPCa may have abnormal BP for several reasons: (1) MRI-invisible cancer not adjacent to the MRI-visible lesion; (2) a field effect of prostate changes possibly related to predisposition to csPCa; and/or (3) reactive changes to the presence of csPCa in part of the gland. The existence of MRI-invisible cancer is well known (26–28), though csPCa is more likely to be near MRI-visible lesions and would be encompassed by our buffer analyses (8). csPCa is also known to arise in the context of a field effect of prostate-wide conditions that predispose to oncogenesis (11). *T*_*2*_ changes in BP might reflect pre-cancerous changes, including inflammation, chemical irritation, pathogen exposure, and/or modifications in local gene expression (29). A recent study described striking genomic alterations of benign prostate tissue in patients with csPCa (12). Future studies should leverage advanced quantitative imaging techniques like luminal water imaging (2) and MR spectroscopy (30) to better assess the *T*_*2*_ properties of BP in patients with and without csPCa. Regardless of etiology, the abnormal BP in patients harboring csPCa suggests the presence of interesting biological processes beyond what is reflected in MRI-visible lesions.

*T*_*2*_-weighted signal in BP contributes meaningful information and may have diagnostic or prognostic clinical value. We have previously shown that a quantitative DWI biomarker called RSI *C*_*1*_ is useful for voxel-level and patient-level detection of csPCa (14,15). Median *T*_*2*_ in the prostate was not correlated with maximum RSI *C*_*1*_, and patient-level csPCa detection improved after normalizing maximum RSI *C*_*1*_ by median *T*_*2*_. These findings suggest *T*_*2*_ signal of the entire prostate, not just of the radiographically visible lesions, may provide insight into the physiological changes linked to csPCa and may improve risk stratification of patients with suspected prostate cancer. As a practical consideration for incorporating this information into clinical practice, we demonstrated that median *T*_*2*_ measurements from the entire prostate (visible lesions included) were largely indistinguishable from median values of the BP alone, with nearly identical csPCa detection performance. Whole-prostate contouring is relatively simple and can now be performed automatically with freely-available AI tools (31), obviating the need for expert delineation of lesions by specialized radiologists in order to integrate BP signal information into diagnostic decision making.

Classification performance differed between Cohort 1 and Cohort 2. This may simply reflect chance variation, but there are some noteworthy differences between the cohorts that reflect dynamic patterns in clinical practice over time. The chronologically more recent Cohort 2 included proportionally more patients with a prior biopsy. Cohort 2 also had more patients with targeted-only biopsy. Perhaps most importantly, when the data from Cohort 2 was being acquired, urologists at our institution had become increasingly reliant on MRI results in deciding which patients to biopsy, such that Cohort 2 had zero participants with PI-RADS 1 lesions, compared to over 20% of Cohort 1. Inclusion of more PI-RADS 1 patients would be expected to improve the performance of MRI, and the AUC for Cohort 1 was indeed higher than for Cohort 2.

A limitation of this study is that proton density weighted data were not acquired for the patients included in this study, so we could not directly examine the proton density of BP. However, since imaging hardware was fixed across all patients, tissue proton density should be the principal determinant of *S*_*0*_. Limitations of retrospective studies also apply. This was a single-center, single-scanner study, though two different acquisition protocols were used. Future studies will also leverage whole-mount histopathology and explore molecular and genomic alterations associated with *T*_*2*_ abnormalities in benign and benign-appearing tissue.

We conclude that the background tissue of the prostate exhibits systematically abnormal *T*_*2*_-weighted MRI in patients who harbor csPCa. This global prostate effect appears complementary to the focal diffusion restriction characteristic of suspicious visible lesions. In sum, MRI signal outside visible lesions may afford untapped diagnostic value for detection of csPCa. These intriguing initial findings should be validated in broader datasets.

## Data Availability

All data produced in the present study are available upon reasonable request to the corresponding author

## Supplemental Materials

**Supplemental Figure 1:**
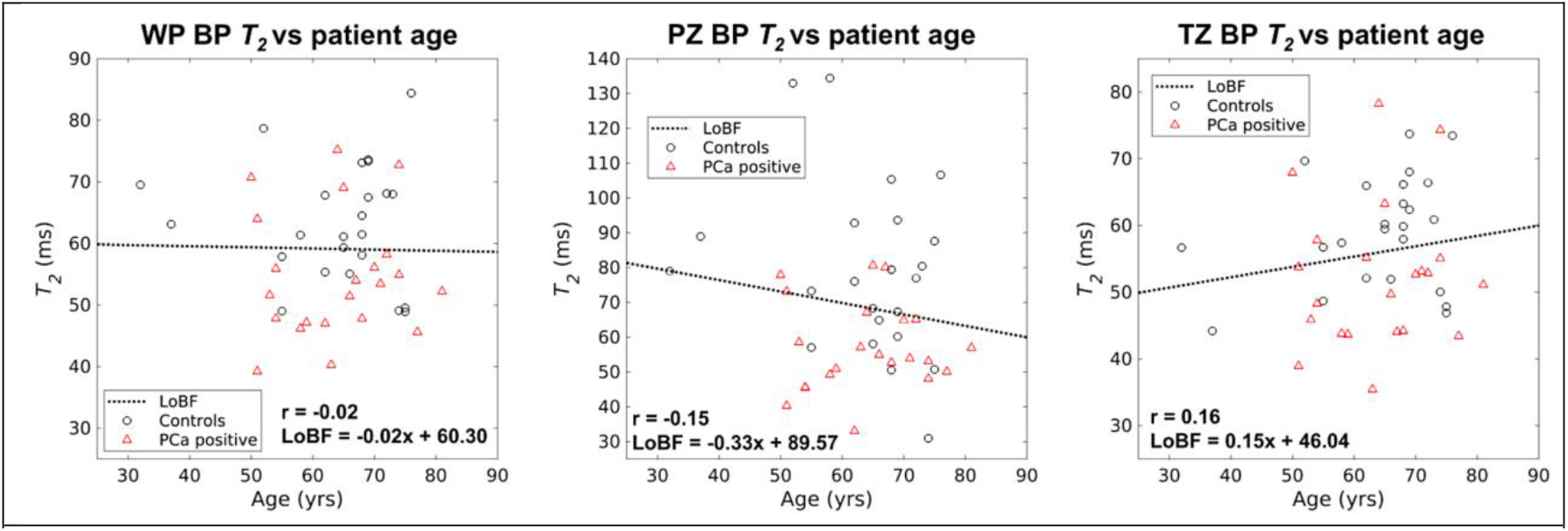
Correlation of *T*_*2*_ time in BP with patient age in cohort 1. *T*_*2*_ was not significantly correlated with patient age in any of the prostatic zones (*p*>0.05 for WP, PZ, and TZ). r: Pearson correlation coefficient, LoBF: line-of-best-fit to the data.

**Supplemental Figure 2:**
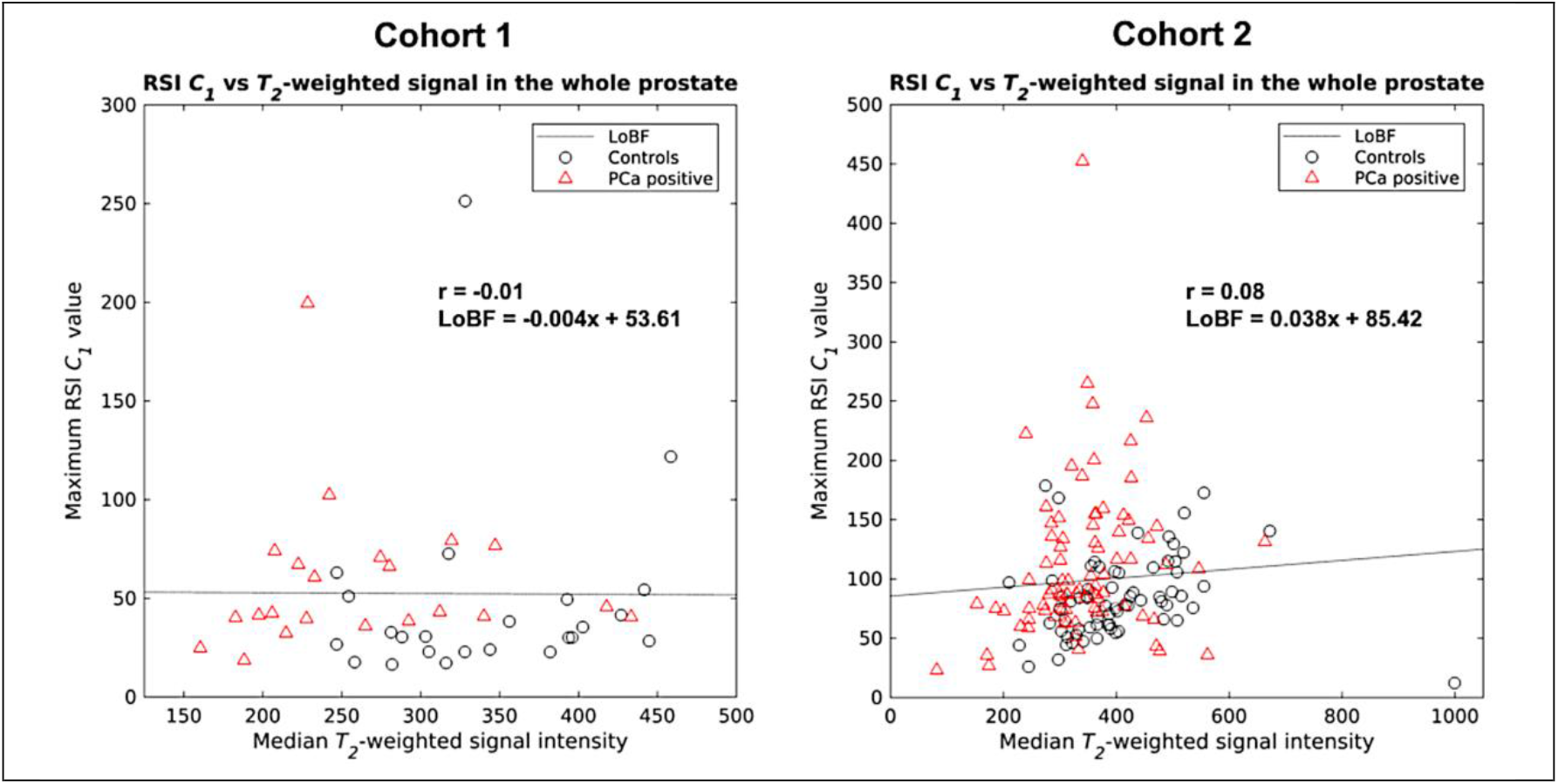
Scatterplots comparing whole-prostate measurements of maximum RSI *C*_*1*_ and median *T*_*2*_-weighted signal intensity. No significant correlation was observed between the two metrics in either cohort (*p*>0.05). r: Pearson correlation coefficient, LoBF: line-of-best-fit to the data.

